# Generation of guideline-based clinical decision trees in oncology using large language models

**DOI:** 10.1101/2024.03.04.24303737

**Authors:** Brenda Y. Miao, Eduardo Rodriguez Almaraz, Amir Ashraf Ganjouei, Arvind Suresh, Travis Zack, Maxim Bravo, Srinidhi Raghavendran, Boris Oskotsky, Ahmed Alaa, Atul J. Butte

## Abstract

**Background:** Molecular biomarkers play a pivotal role in the diagnosis and treatment of oncologic diseases but staying updated with the latest guidelines and research can be challenging for healthcare professionals and patients. Large Language Models (LLMs), such as MedPalm-2 and GPT-4, have emerged as potential tools to streamline biomedical information extraction, but their ability to summarize molecular biomarkers for oncologic disease subtyping remains unclear. Auto-generation of clinical nomograms from text guidelines could illustrate a new type of utility for LLMs.

**Methods:** In this cross-sectional study, two LLMs, GPT-4 and Claude-2, were assessed for their ability to generate decision trees for molecular subtyping of oncologic diseases with and without expert-curated guidelines. Clinical evaluators assessed the accuracy of biomarker and cancer subtype generation, as well as validity of molecular subtyping decision trees across five cancer types: colorectal cancer, invasive ductal carcinoma, acute myeloid leukemia, diffuse large B-cell lymphoma, and diffuse glioma.

**Results:** Both GPT-4 and Claude-2 “off the shelf” successfully produced clinical decision trees that contained valid instances of biomarkers and disease subtypes. Overall, GPT-4 and Claude-2 showed limited improvement in the accuracy of decision tree generation when guideline text was added. A Streamlit dashboard was developed for interactive exploration of subtyping trees generated for other oncologic diseases.

**Conclusion:** This study demonstrates the potential of LLMs like GPT-4 and Claude-2 in aiding the summarization of molecular diagnostic guidelines in oncology. While effective in certain aspects, their performance highlights the need for careful interpretation, especially in zero-shot settings. Future research should focus on enhancing these models for more nuanced and probabilistic interpretations in clinical decision-making. The developed tools and methodologies present a promising avenue for expanding LLM applications in various medical specialties.

**Key Points:**

- Large language models, such as GPT-4 and Claude-2, can generate clinical decision trees that summarize best-practice guidelines in oncology
- Providing guidelines in the prompt query improves the accuracy of oncology biomarker and cancer subtype information extraction
- However, providing guidelines in zero-shot settings does not significantly improve generation of clinical decision trees for either GPT-4 or Claude-2

## Introduction

Molecular biomarkers are becoming increasingly crucial in supporting the diagnosis and treatment of oncologic diseases but keeping up with the latest guidelines and relevant research can be time-consuming for physicians, researchers, and patients. The recent emergence of several new large language models (LLMs) present a unique opportunity to help streamline text-heavy healthcare workflows, including medical information summarization and education. Previous studies have demonstrated that new LLMs are capable of extracting complex clinical information from oncology progress notes^1^, suggesting differential diagnoses^2^, or even generating decision trees from clinical trial criteria^3^ or for clinical decision support^4^. The generation of decision trees can provide clear visual guidelines for clinical support, which can significantly impact downstream clinical care. In this study, we aimed to assess the capabilities of two recently developed LLMs in generating diagnostic decision trees for the molecular subtyping of cancers, using published clinical guidelines.

## Methods

Diagnostic trees describing cancer subtypes based on molecular biomarker status were generated for five cancers using GPT-4 (OpenAI) and Claude-2 (Anthropic), two LLMs with public Application Programming Interfaces (APIs). These cancers were selected based on the prevalence of known molecular biomarkers, and included two common solid organ cancers (colorectal cancer [CRC] and invasive ductal carcinoma [IDC]), a common hematologic cancer (acute myeloid leukemia, AML), a rare hematologic cancer (diffuse large B-cell lymphoma [DLBCL]), and a rare solid cancer (diffuse glioma).

Trees were generated using a specific prompt that contained either only formatting guidelines (Figure 1) or also included information provided from recent classification guidelines for each of the five cancers^5–9^ (Table S2). Clinical trees were generated to contain molecular biomarker status as nodes, terminating at nodes that were molecular subtypes. Model temperature was set to 0, and a new API call was made for each of the different prompts used. Additional details on models and parameters used are provided in Supplemental Table 1. Results were processed into Pydot graph objects^10^ and visualized using an interactive dashboard developed using Streamlit^11^.

**Figure 1.**
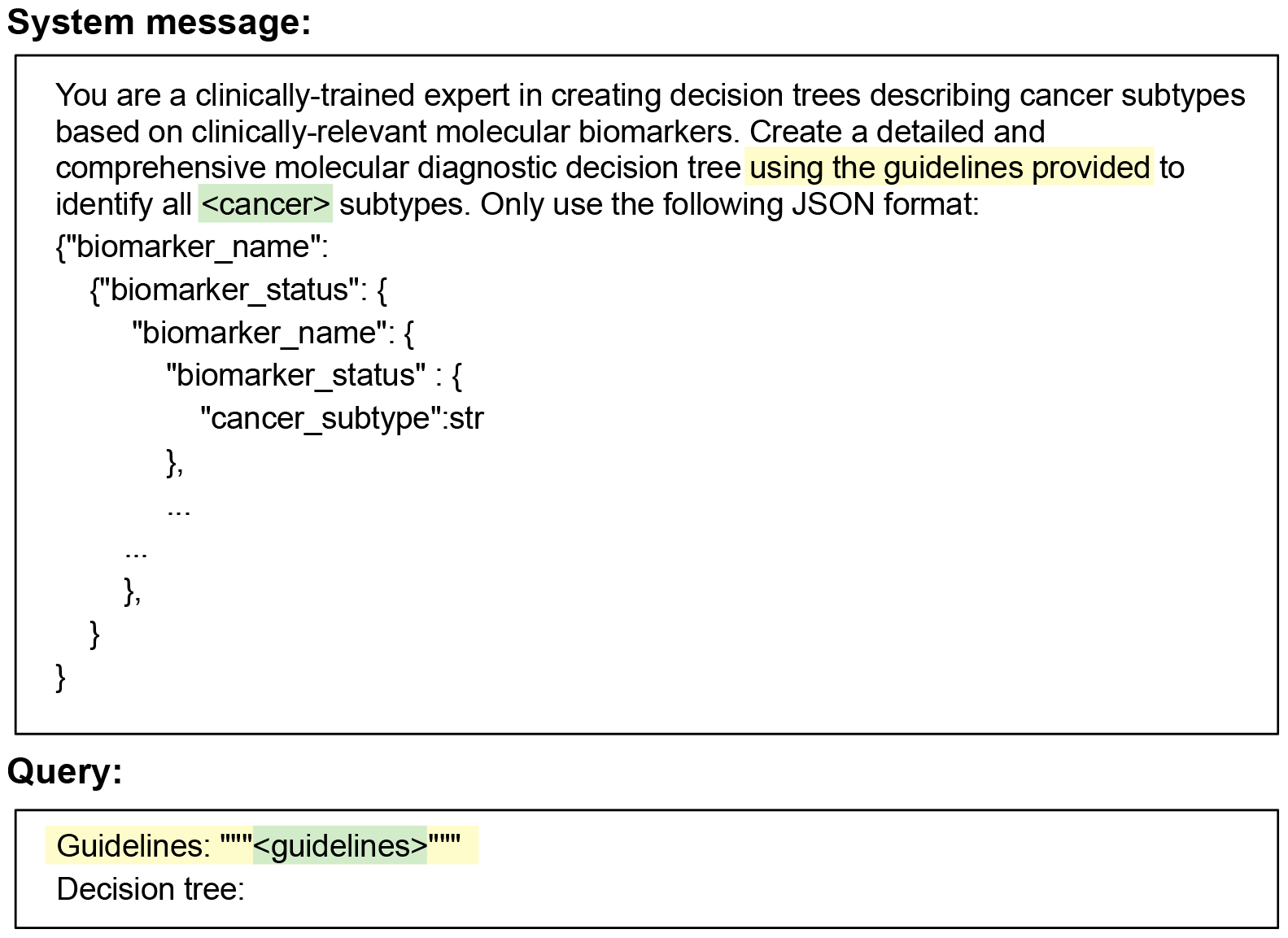
Prompts to generate clinical decision trees. Prompt used to generate clinical cancer subtyping trees. Values highlighted in green are replaced with cancer specific information for each of the five cancers evaluated, and values highlighted in yellow are only included if guidelines are present.

**Figure 2.**
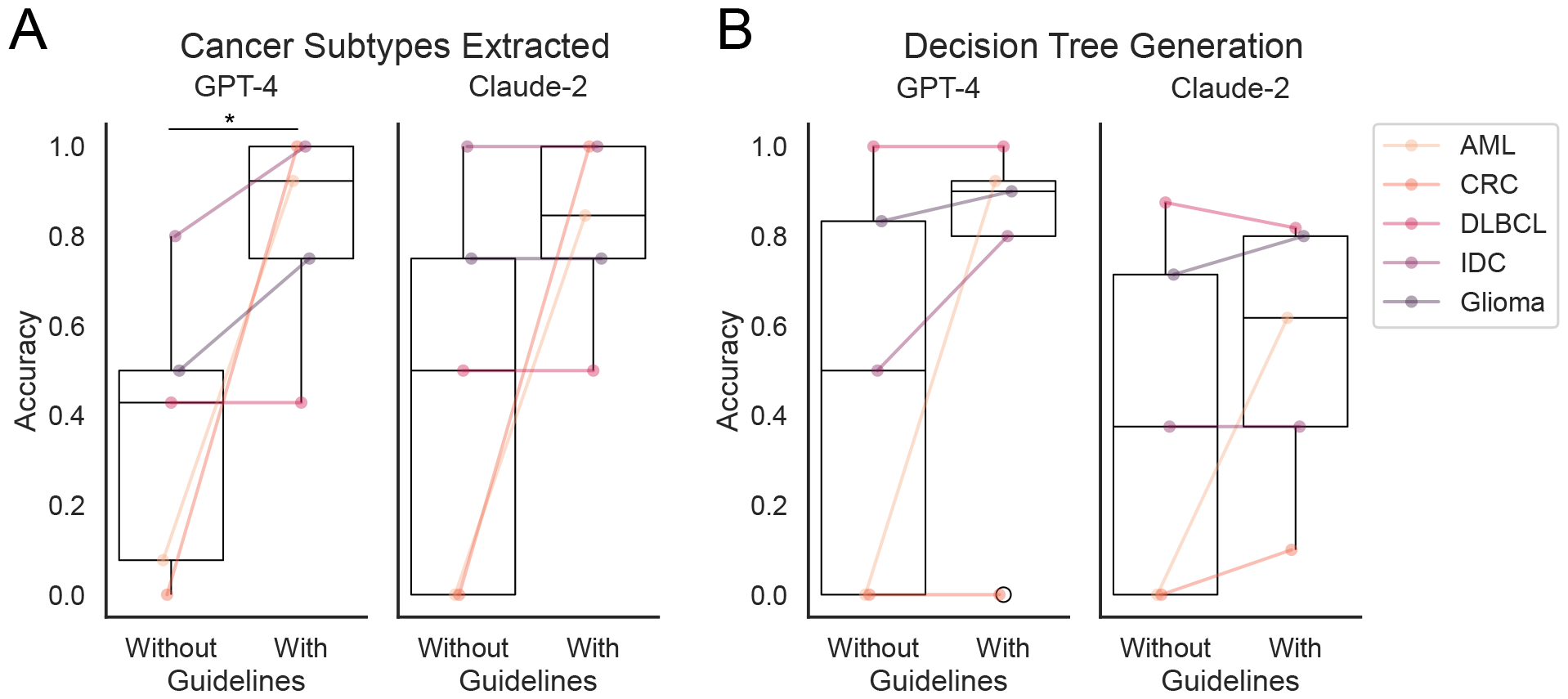
Accuracy of clinical decision tree generation using LLMs. Clinical evaluators assessed the A) accuracy of cancer subtype extracted by each LLM with and without guidelines. B) Clinical evaluators also assessed the overall accuracy of clinical decision trees generated. A tree was only considered correct if all biomarkers and subtypes were clinically appropriate, and the biomarkers accurately described the associated cancer subtype.

Each branch of LLM-generated decision trees were evaluated against subtyping decision trees generated by clinical reviewers based on clinical guidelines. Evaluators were blinded to which language model generated which tree, and each tree was evaluated by two reviewers, with discrepancies resolved by discussion. We report mean accuracies of subtyping trees, as well as proportions of subtypes and biomarkers correctly extracted by the two LLMs for each cancer. Hallucinations, identified as values not mentioned in recent guidelines for use in molecular cancer subtype diagnosis, were also quantified by clinical evaluators. Accuracy of LLM trees with and without guidelines were compared with two-sided T-tests using Scipy^12^. A p-value less than 0.05 was considered statistically significant.

## Results

Both Claude-2 and GPT-4 were able to create properly formatted decision trees with or without being provided actual clinical guideline text. Including guideline text improved the proportion of cancer subtypes and biomarkers that each model was able to extract. Mean accuracy of cancer subtype extraction increased when guidelines were provided, with the Claude-2 model increasing from 45% (SD: 44.7%, n=5) to 81.9% (SD: 20.8%, p=0.13) and GPT-4 from 36.1% (SD: 33.3%) to 82.0% (SD: 24.2%, p=0.035). Without guidelines, both GPT-4 and Claude-2 were best at generating accurate cancer subtypes in decision trees for IDC (80% and 100%, respectively) and neither were able to produce subtypes of CRC. By providing guideline text, both GPT-4 and Claude-2 were able to extract and visualize all expected subtypes for IDC and CRC (Figure S1A).

Regarding hallucinations, GPT-4 and Claude-2 produced the greatest proportion of hallucinated subtypes, which were subtypes not present in clinical trees generated by clinical annotators, for CRC and AML when not provided guideline text. Subtypes that were not mentioned in recent guidelines, such as “NPM1 Wildtype, FLT3-ITD Wildtype and CEBPA Mutated AML,” were considered hallucinations. On average, 40% (SD: 54.8%) of subtypes extracted by Claude-2 without guidelines were deemed to be hallucinations, which decreased to 21.0% (SD: 23.7%, p=0.50) when provided guideline text. GPT-4 referenced hallucinated cancer subtypes 37.1% (SD: 54.8%) of the time when not provided guideline text, which dropped to 2.9% (SD: 6.3%, p=0.17) when provided with guideline text (Figure S1B).

For accurate biomarker extraction, Claude-2 extracted 55.3% of expected biomarkers on average (SD: 24.6%) without guideline text and 86.2% with (SD: 16.4%, p=0.07), while GPT-4 extracted 50.3% (SD: 27.3%) of biomarkers without guideline text and 83.3% with (SD: 23.5%, p=0.048). Without guideline text, both GPT-4 and Claude-2 both showed 75% accuracy for biomarker extraction for IDC and were least accurate in extracting biomarkers for AML (4.2% and 12.5%, respectively). With guideline text, both GPT-4 and Claude-2 were able to extract all expected subtypes for IDC and diffuse gliomas (Figure S2A).

On average, without guideline text, Claude-2 and GPT-4 produced biomarkers that were considered hallucinations (for example, “RBM15::MKL1” and “TP53”) in 16.3% (SD: 17.1%) and 16.0% (SD: 35.8%) of generated values, respectively. With provided guideline text, hallucinations decreased to 12.5% (SD: 13.6%) for Claude-2 and 13.0% (SD: 15.9%) for GPT-4. The largest proportion of hallucinated biomarkers was produced for AML, with 40% hallucinations for Claude-2 and 80% for GPT-4, although providing guidelines reduced model hallucination down to 8.7% for Claude-2 and 7.7% for GPT-4 (Figure S2B).

Assessment of average overall accuracy of decision trees showed that, without guidelines, GPT-4 produced valid branches 46.7% (SD: 46.2%) of the time, while decision tree branches were 39.3% (SD: 40.1%) valid for Claude-2. Substantial increases in decision tree accuracy were seen for AML, going from 0% to 92.3% for GPT-4 and 0% to 61.7% for Claude-2. However, adding in guideline text did not significantly increase overall accuracy of decision tree generation for either GPT-4, which increased to 72.5% (SD: 41.1%, p=0.38) or Claude-2 (54.2%, SD: 30.5%, p=0.52).

A streamlit dashboard was developed to provide a user interface for exploration of GPT-4 and Claude-2 model performance on subtyping tree extraction for user-specified cancer types and guidelines (Figure 3).

**Figure 3.**
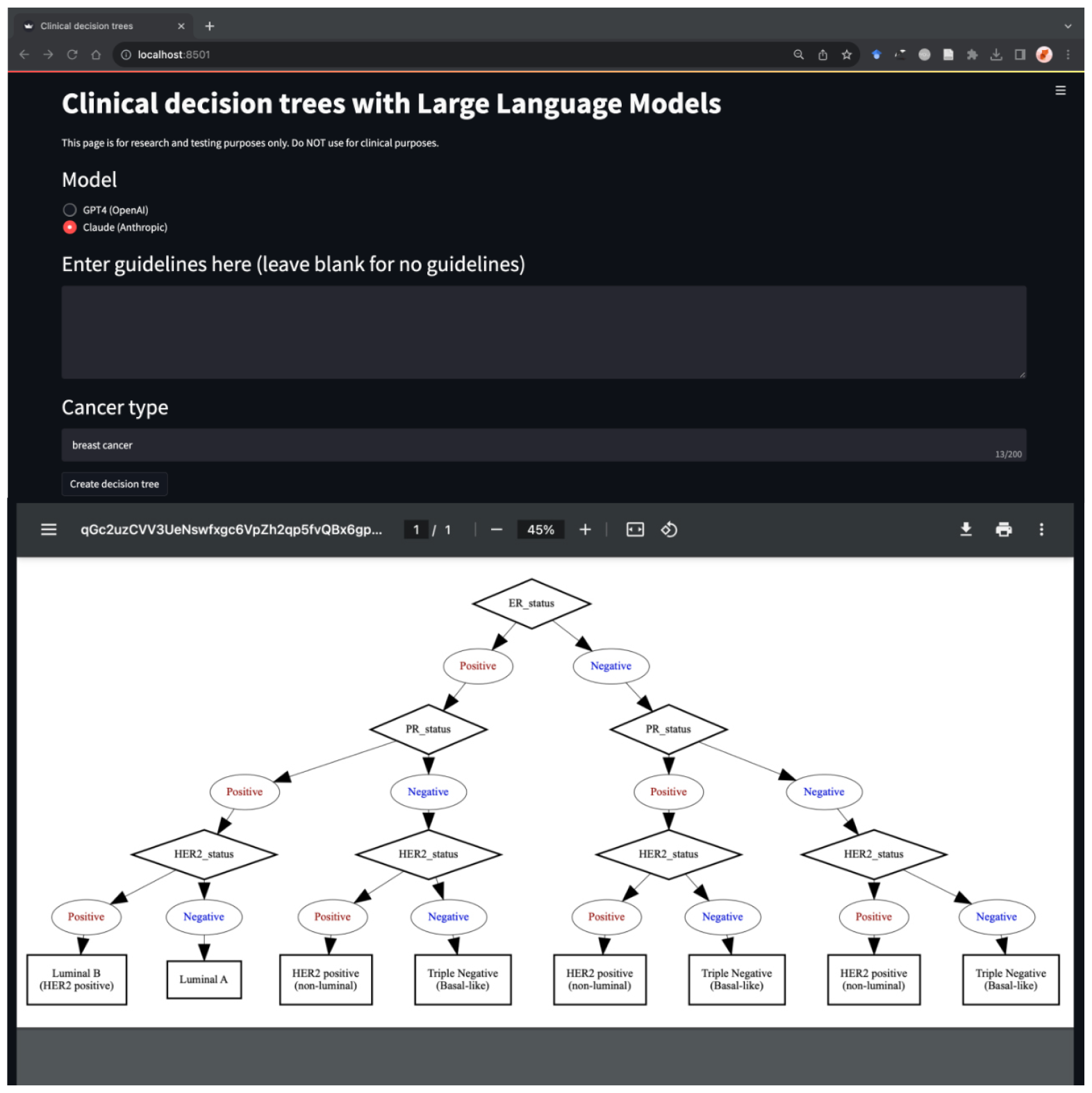
Clinical decision tree dashboard. A streamlit dashboard was created to enable exploration of subtyping decision trees for other cancers and guidelines. The dashboard can be found at https://clinicaltrees.org/

## Discussion

Here, we demonstrate the capability for language models to generate accurate and comprehensive decision trees from clinical guideline text for molecular diagnosis across multiple cancer types. Additionally, we showed that adding clinical guideline text into prompts improves extraction of molecular biomarkers and oncology disease subtypes, but did not significantly improve clinical decision tree generation.

While this brief report identifies opportunities for LLMs in supporting biomedical information review and visualization in oncology, the results are focused on molecular diagnosis, which is only a part of clinical decision making. Furthermore, not all molecular features are binary in nature, and future iterations of these decision trees may be assessed for their ability to include probabilities at each branch along the decision tree. Finally, another limitation to this study is the use of API-based models, which are not as interpretable and are more costly to run compared to open-source alternatives. We also did not perform any prompt engineering, and further exploration of strategies like chain-of-thought may help improve decision tree generation, which involves significant reasoning capabilities.

Despite these limitations, our initial evaluation of GPT4 for oncology molecular information extraction shows significant potential for further development. Additionally, we provide open access to the tools assessed and developed here, and for future studies to use similar approaches to evaluate summarization of guidelines for treatment or other aspects of clinical workflows across different medical specialties. Future work might even include being able to summarize many raw clinical studies and results from clinical trials into more accessible guideline texts and visualizations.

## Data availability

All prompts and text guidelines used are referenced or provided in supplemental materials. All code used to generate and visualize the clinical trees can be found at https://github.com/BMiao10/clincial-decision-trees. The streamlit dashboard for exploration of decision trees for other values can be found at https://clinicaltrees.org/

## Supplemental figures

**Supplemental table 1.**
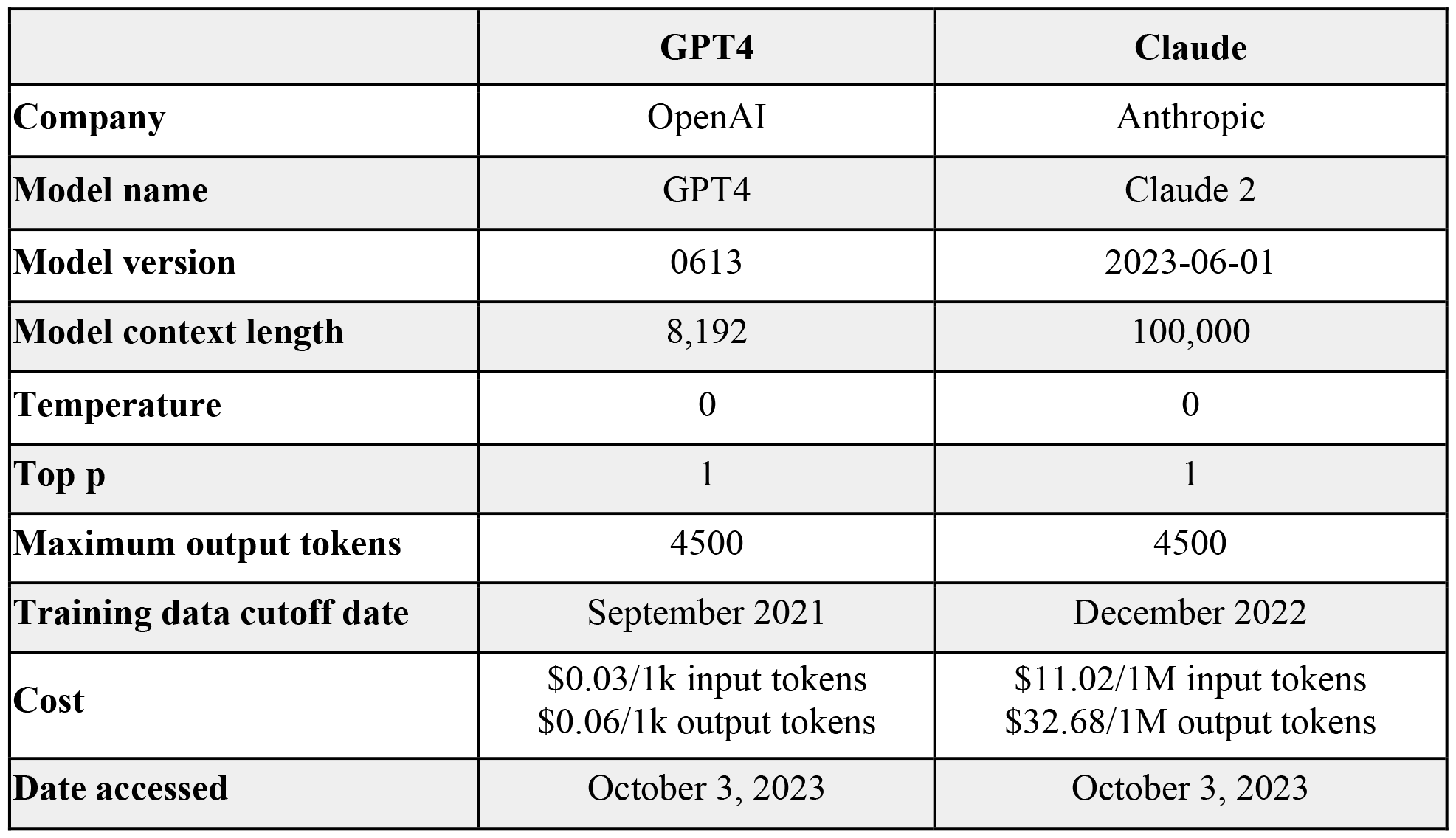
Language model overview. Overview of language models used, context length, date accessed, training data cutoff, and parameters used.

|  | **GPT4** | **Claude** |
| --- | --- | --- |
| **Company** | OpenAI | Anthropic |
| **Model name** | GPT4 | Claude 2 |
| **Model version** | 0613 | 2023-06-01 |
| **Model context length** | 8,192 | 100,000 |
| **Temperature** | 0 | 0 |
| **Top p** | 1 | 1 |
| **Maximum output tokens** | 4500 | 4500 |
| **Training data cutoff date** | September 2021 | December 2022 |
| **Cost** | \$0.03/1k input tokens<br>\$0.06/1k output tokens | \$11.02/1M input tokens<br>\$32.68/1M output tokens |
| **Date accessed** | October 3, 2023 | October 3, 2023 |

**Supplemental table 2.**
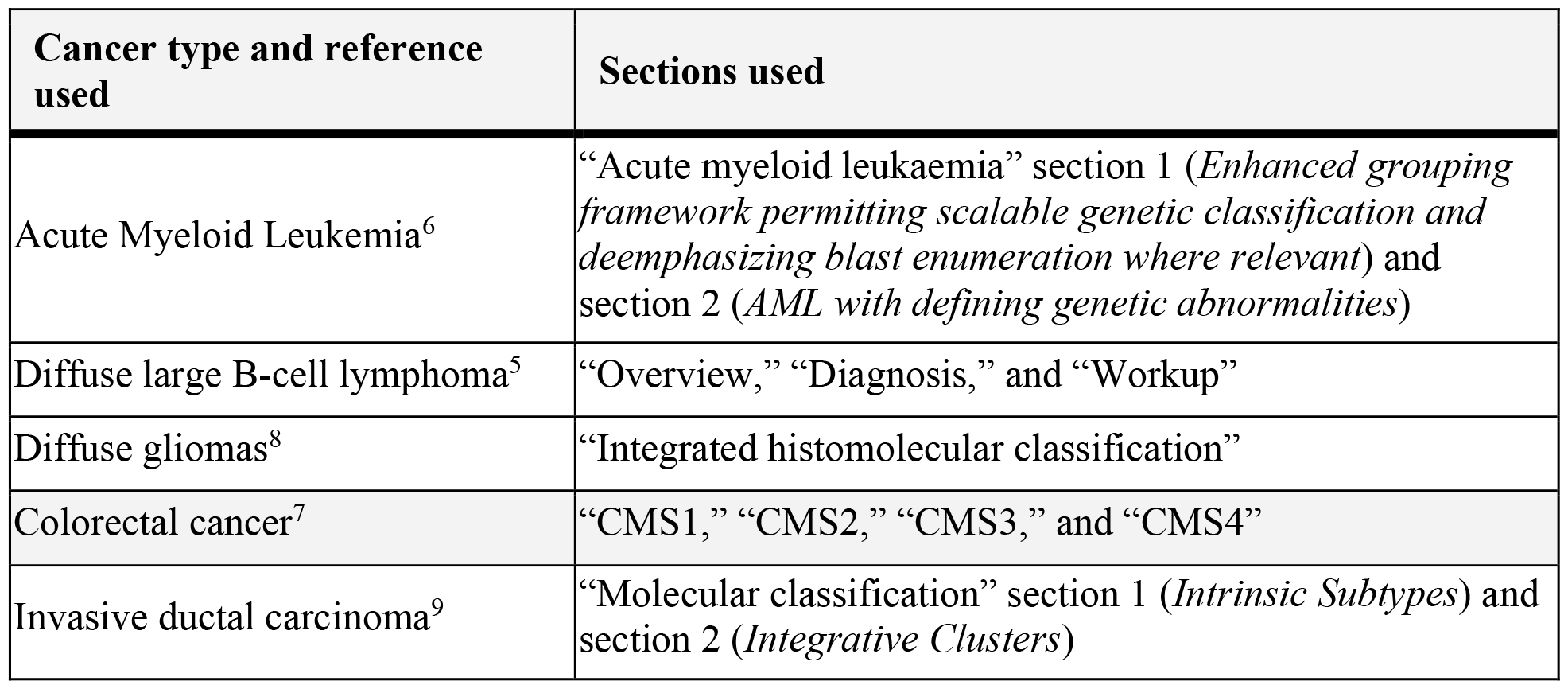
Guideline references and sections used.

**Supplemental figure 1.**
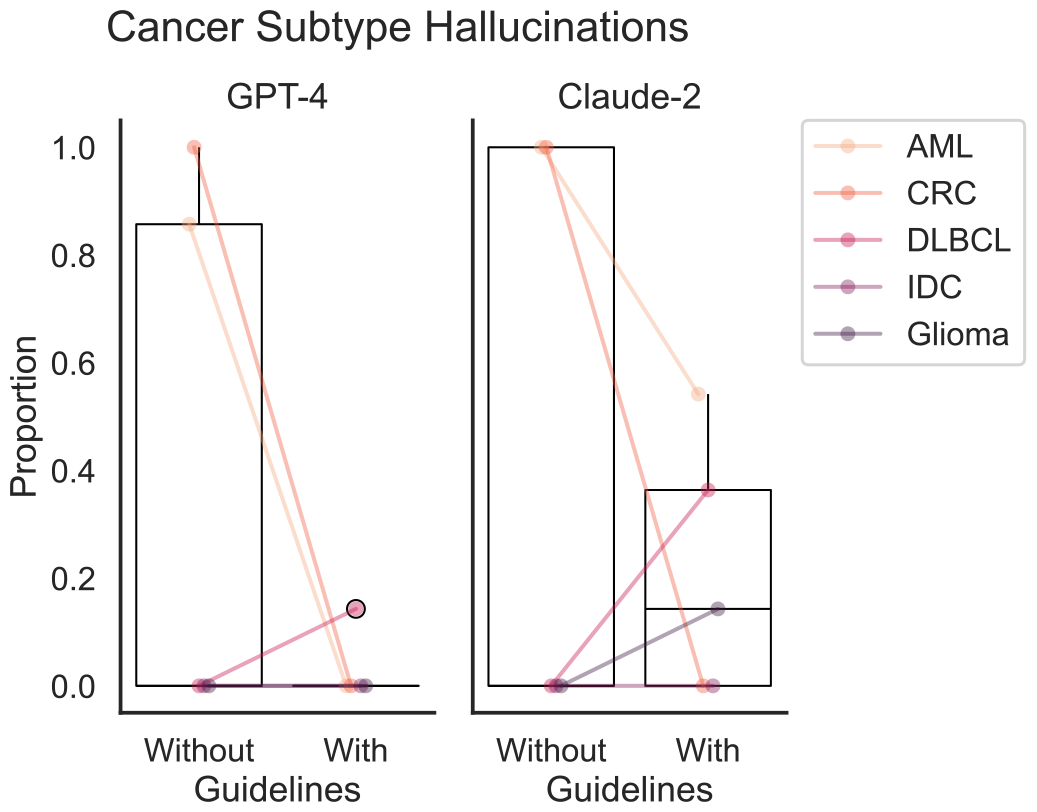
Accuracy and hallucinations of biomarker extraction.

**Supplemental figure 2.**
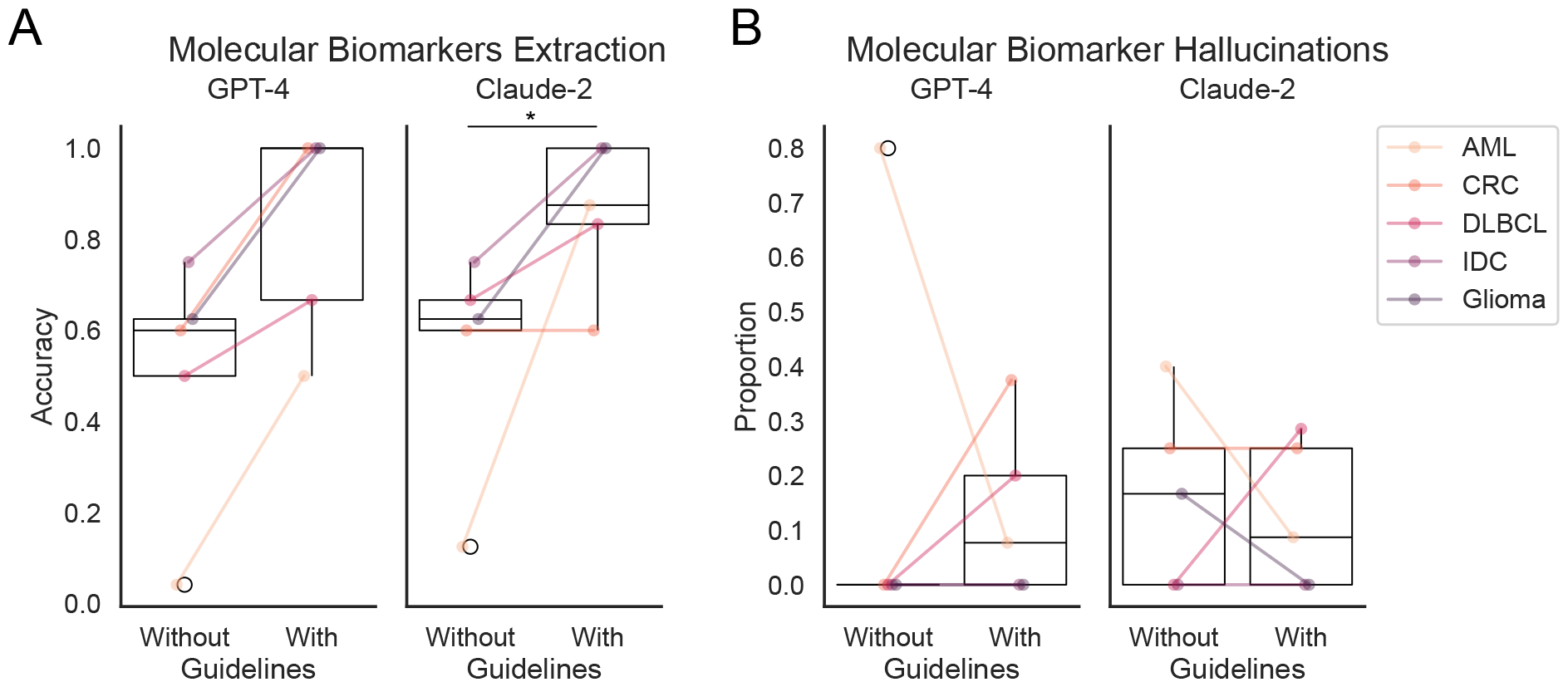
Accuracy and hallucinations of cancer subtype extraction.

